# COVID-19 pandemic: every day feels like a weekday to most

**DOI:** 10.1101/2020.05.11.20098228

**Authors:** Tony Liu, Jonah Meyerhoff, David C. Mohr, Lyle H. Ungar, Konrad P. Kording

## Abstract

The COVID-19 outbreak has clear clinical^1^ and economic^2^ impacts, but also affects behaviors e.g. through social distancing^3^, and may increase stress and anxiety. However, while case numbers are tracked daily^4^, we know little about the psychological effects of the outbreak on individuals in the moment. Here we examine the psychological and behavioral shifts over the initial stages of the outbreak in the United States in an observational longitudinal study. Through GPS phone data we find that homestay is increasing, while being at work dropped precipitously. Using regular real-time experiential surveys, we observe an overall increase in stress and mood levels which is similar in size to the weekend vs. weekday differences. As there is a significant difference between weekday and weekend mood and stress levels, this is an important decrease in wellbeing. For some, especially those affected by job loss, the mental health impact is severe.

## Introduction

The ongoing COVID-19 outbreak has become a global crisis, not just because of rising infection and death rates but also because of the behavioral changes it necessitates. Even those who are not directly affected by the disease have seen their lives disrupted through mandated social distancing measures. These sudden changes can impact an individual’s mental wellbeing: reduced social connections is linked to depression^5^, while physical restrictions of quarantine can also have negative psychological effects^6^. In addition to the outbreak’s clinical and economic impact, we also need to understand these mental wellness and behavioral changes in order to combat the disease.

Both wellbeing and behavior can be measured digitally, such as through online surveys^7^, or location and survey data collected by phone sensors^8,9^. There has already been work examining the spread of COVID-19 in China using mobility data from the social media platform Tencent^10^ as well as work measuring mental health-related effects of the outbreak through cross-sectional web surveys^11,12^. However, only ecological momentary assessments (EMAs) allow us to understand how the situation affects the way we feel in the moment. In this article we explore how people are changing their behavior in the first weeks of the pandemic, and the nature of the associated changes in wellbeing and psychological distress as measured with EMAs.

We use data on 127 U.S. adult participants (Table 1) from an ongoing longitudinal study, where they completed baseline assessments of their mental wellbeing and installed a personal sensing application on their phones. This application administers EMAs asking questions about their mood (“How is your mood?”), stress (“Are you stressed?”), rated on 9-point Likert scales in the morning, afternoon and evening as participants go about their daily lives, allowing us to track day-to-day changes in wellbeing. The PHQ-8^9^ is administered at 3-week intervals to measure depressive symptom severity. The app also records detected GPS locations, and asks questions about the locations participants have visited (“What kind of place is this?”). From GPS data, we measure how much time participants spent at work and at home, as well as their general movement levels as the outbreak unfolds. We then use these location measures with EMA data to compare our participant’s behavior and mental wellbeing in study weeks before and after the national state of emergency declaration (3/13)^13^ social distancing guidelines (3/16)^14^.

**Table 1:** Demographic summary of participants (n=127).

| **<u>Demographics</u>** |
| --- |
| Age: mean=42.4 years, std=12.0 |
| Race: White 68.5%, African American 22.8%, More than one race 5.5%, Asian 2.4% |
| Gender: 70% female |
| Previous major depressive disorder diagnosis: 54.3% |

## Results

Location and EMA Phone Sensors pre-COVID-19 mandates (3/3-3/9) and post-COVID-19 mandates (3/24-3/30)

We want to understand shifts in behavior and wellbeing that coincide with mandated social distancing and other quarantine measures. We saw sharp decreases in time spent at work (Cohen’s d=0.33, 95% CI [-5.26, −2.12], p<0.001) as well as increases in time spent at home (Cohen’s d=0.38, 95% CI [7.07, 13.83], p<0.001) that coincided with governmental mandates of social distancing (Figure 1, Table 2). The corresponding increases in stress (Cohen’s d=0.15, 95% CI [0.03, 0.41], p=0.03) and mood (Cohen’s d=0.17, 95% CI [0.08, 0.41], p<0.01) across the two weeks are also statistically significant. This can be compared to the weekday/weekend distinction of stress (Cohen’s d=0.23, 95% CI [0.19, 0.45], p<0.01) and mood (Cohen’s d=0.13, 95% CI [0.06, 0.30], p<0.01) during the pre-COVID period.

**Figure 1.**
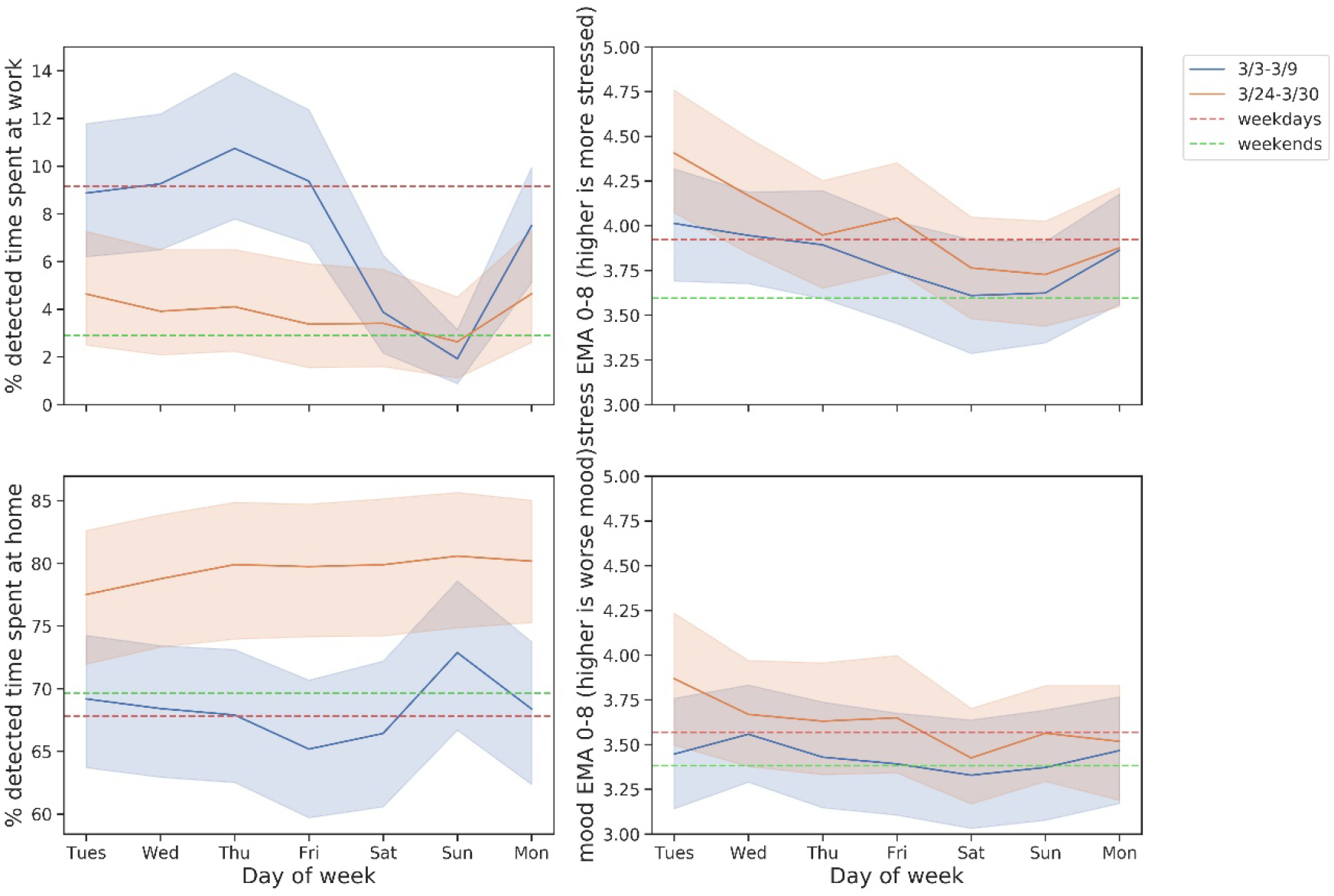

**Table 2:** Full hypothesis testing information for the paired two-tailec survey data. Reported p-values are uncorrected.

| **comparison** | **sensor/survey** | **t-statistic** | **dof** | **p-value** | **95% CI** | **Cohen's d** |
| --- | --- | --- | --- | --- | --- | --- |
| pre/post COVID guidelines | Work percentage | -4.645 | 119 | 8.843e-06 | [-5.26, -2.12] | 0.330 |
| pre/post COVID guidelines | Home percentage | 6.119 | 119 | 1.242e-08 | [7.07, 13.83] | 0.382 |
| pre/post COVID guidelines | PHQ-8 | 1.665 | 112 | 0.0987 | [-0.11, 1.29] | 0.106 |
| pre/post COVID guidelines | Stress EMA | 2.256 | 126 | 0.0258 | [0.03, 0.41] | 0.148 |
| weekdays/weekends | Stress EMA | 4.789 | 125 | 4.645e-06 | [0.19, 0.45] | 0.231 |
| pre/post COVID guidelines | Mood EMA | 2.871 | 126 | 0.00480 | [0.08, 0.41] | 0.165 |
| weekdays/weekends | Mood EMA | 2.936 | 125 | 0.00395 | [0.03, 0.30] | 0.131 |

For EMA-administered PHQ, we find that the changes within individuals across the two study periods were not significant (Cohen’s d=0.11, 95% CI [-0.11, 1.29], p=0.1). However, we see that individuals who have been directly impacted by the crisis through job loss show worsening of depressive symptoms. Seven individuals reported job loss during the post-social distancing study week, and all seven also reported an increase in their depression severity (median increase of 3 points); the probability of that happening by chance is 1/128 (Table 3). These results are consistent with other findings^15^ that show the direct impact of the virus on mental wellbeing through income instability is severe.

**Table 3:** Changes in PHQ-8 survey scores from study week 3/3-3/9 to study week 3/24-3/30. PHQ-8 scores range from 0 to 24, with higher values indicating more severe depressive symptoms.

| **3/24-3/30 reported job loss** | **n** | **mean** | **std** | **min** | **25%** | **50%** | **75%** | **max** |
| --- | --- | --- | --- | --- | --- | --- | --- | --- |
| no | 106 | 0.417 | 3.812 | -12 | -1.5 | 0 | 2 | 18 |
| yes | 7 | 3.214 | 1.380 | 1.5 | 2.25 | 3 | 4.25 | 5 |

## Discussion

We did not see evidence of significant worsening in depression at a population level in the first weeks following the national declaration of emergency and social distancing guidelines. We did see significant increases in mood and stress; however these were small. At a population level, the stress and mood impact of the outbreak is comparable to every day being a weekday. Our results suggest a significant increase in depressive symptom severity for those affected by job loss. Though our findings are not consistent with the kind of massive mental health crisis at a population level that some in the media speculate them to be, our data do suggest that the mental health consequences are likely concentrated in groups that have been substantially affected by the pandemic, such as those who have lost employment. There are undoubtedly other groups that experience strong influences, and further investigation into these at-risk groups is needed to understand the nature of any psychological impact so that mental health services can be appropriately allocated for the crisis.

It is important to note the sampling limitations of our study. Our participants are recruited from a research panel and are not wholly representative of national demographics. In particular, our sample over-represents females and individuals with previously diagnosed depression (Table 1). We also note that our analysis is based on completed EMAs for each individual and does not account for missed surveys. The survey response rate dropped in the week post declaration of emergency compared to previous weeks (75% response rate to 65% response rate), so it is possible that the participants most affected by the crisis are self-censoring^16^. Furthermore, as we only look at short-term effects of the coronavirus crisis, these findings may change as time goes on. We will continue monitoring the longer-term response to the crisis and will report on trends as they emerge. Although it remains to be seen how individuals respond as the coronavirus crisis progresses, these findings suggest that at the population level and on a short time scale, the COVID-19 pandemic has increased stress levels widely, but has not had a uniform impact on mental health. Rather, it is likely concentrated in at-risk groups such as those who have lost employment.

## Methods

### Participants

Participants across the United States were invited to participate in our study through Focus Pointe Global, a national research panel, in an enrollment period beginning February 4, 2020 and ending February 10, 2020. We included participants who were at least 18 years old, owned an Android phone, and who did not report any prior diagnosis of severe mental illness (e.g. bipolar disorder, psychotic disorder). Participant demographic characteristics can be found in Table 1. All study protocols were approved by Northwestern University’s Institutional Review Board.

### Data Collection and Processing

Participation in the 16 week study which began February 11, 2020 included completing online surveys administered via REDCap^17,18^ at baseline and at check-ins occurring once every three weeks. Participants also installed Passive Data Kit (PDK)^12^, a library and mobile app for collecting cell phone sensor data, on their phones. Sensor data collected include real-time GPS coordinates as well as EMA surveys administered three times a day for seven consecutive “check-in” days that occur every three weeks.

When participants were in a check-in week, they were prompted via phone notification at their preferred times of day. The stress (“are you stressed?”) and mood (“how is your mood?”) EMAs were administered at all three daily check-ins with a nine-point Likert scale. The evening survey also presented a map and prompted participants to label particular stationary locations they were detected to have visited throughout the day (“what kind of place is this?). At the beginning and end of the check-in week, an additional set of EMA questions were sent to the participants which correspond to the PHQ-8 depression inventory.

The raw GPS data collected by PDK was sampled at roughly 15 second intervals and was transformed into higher-level features of semantic location duration for our study. We assigned a label of “home” or “work” for every GPS reading if it was within 500 meters of the participant-labelled home or work locations. These labelled location readings were then aggregated to produce daily estimates of time spent in both location categories.

### Statistical Analysis

We ran paired, two-tailed Student t-tests of within-individual sensor data in the two most recent check-in weeks of our study: 3/3/20-3/9/20, and 3/24/20-3/30/20. These dates straddle the most prominent government-mandated disruptions with the national declaration of emergency (3/13) and the White House recommendation limiting gatherings (3/16). Daily measurements of semantic location, EMAs, and PHQ were averaged within each week of analysis. Weekend and weekday averages for individuals were calculated over all weeks of our study up through 3/9/20, to align with the last EMA period before coronavirus-related disruptions. The paired values for every individual who had at least one reading in both study periods were used for our t-test, which was 120 participants for location sensors, 127 participants for EMAs, and 113 participants for PHQ-8 surveys. Complete hypothesis testing information can be found in Table 2. Summary statistics on PHQ-8 surveys among participants who recently experienced job loss can be found in Table 3.

### Data Availability

The data are not publicly available due to them containing information that could compromise research participant privacy and consent, but derived and de-identified self-report data will be made available through the NIMH Data Archive at the conclusion of the study.

### Code Availability

The source code for data preparation and analysis is available upon request to the corresponding author TL. The unprocessed source data are not publicly available because of the restrictions noted above.

### Extended Data Legends

Figure 1: Mean work time percentage, home time percentage, mood EMAs, and stress EMAs across two weeks of our study, before and after the national state of emergency declaration. The average sensor values for weekdays and weekends are represented by dotted lines. Shaded regions are bootstrapped 95% confidence intervals.

## End Notes

### Author Contributions

TL performed data cleaning, data analysis, and prepared the manuscript. JM helped write the manuscript. LU, DM, and KK jointly supervised this work and helped write the manuscript. All authors discussed the results and implications throughout the development of the manuscript.

### Materials & Correspondence

Material requests and correspondence should be directed to TL.

